# A conformational variant of p53 (U-p53^AZ^) as blood-based biomarker for the prediction of the onset of symptomatic Alzheimer’s disease

**DOI:** 10.1101/2021.08.23.21261848

**Authors:** Simona Piccirella, Leander Van Neste, Christopher Fowler, Colin L Masters, Jurgen Fripp, James D. Doecke, Chengjie Xiong, Daniela Uberti, Paul Kinnon

## Abstract

**Background:** Ongoing research seeks to identify blood-based biomarkers able to predict the onset and progression of Alzheimer’s disease (AD). A potential biomarker is the unfolded conformational variant of p53, previously observed in individuals in the prodromal and clinical AD stages. In this retrospective study, we compare diagnostic and prognostic performances of measures of the amyloid β load with those of a conformational variant of U-p53 in plasma samples from individuals participating in the Australian Imaging, Biomarkers and Lifestyle (AIBL) cohort.

**Methods:** Immunoprecipitation (IP) followed by liquid chromatography (LC) tandem mass spectrometry (MS/MS) and protein sequencing in plasma samples from the AIBL study identified the clinically relevant AZ 284^®^ peptide, representing a measure of the U-p53 conformational variant (U-p53^AZ^). Based on U-p53^AZ^ quantification via IP/LC electrospray ionisation-coupled MS/MS (AlzoSure^®^ Predict test) on 515 samples from 482 individuals from the AIBL cohort, the predictive performance of U-p53^AZ^ was assessed and compared with amyloid load as measured by amyloid β-positron emission tomography (Aβ-PET). Its predictive performance was determined at 36, 54, 72 and 90 months following baseline assessment.

**Results:** U-p53^AZ^ was able to identify individuals with AD dementia with an area under the receiver operating characteristic curve (AUC) of 99%. U-p53^AZ^ outperformed the conventional Aβ-PET measures in predicting the onset of AD dementia both from preclinical (AUC=98%) and prodromal stages (AUC=89%), even 90 months prior to onset (AUC=99%). Additionally, the estimated predictive performance of U-p53^AZ^ was superior (AUC ≥98%) to other risk factors (i.e., gender, Aβ-PET and *APOE* ε4 allele status) in identifying individuals at high risk for progression to AD.

**Conclusion:** These findings support use of U-p53^AZ^ as blood-based biomarker predicting if individuals, at both asymptomatic and MCI stages, would progress to AD at least six years prior to the onset of clinical AD dementia.

## Introduction

Alzheimer’s disease (AD) is a progressive neurodegenerative disorder, starting with a preclinical phase of normal cognition lasting approximately two decades. While some individuals experience subjective memory complaints (SMC), all eventually progress to mild cognitive impairment (MCI) during the prodromal stage before finally reaching a formal diagnosis of AD dementia (Albert et al., 2011; Aisen et al., 2017). Currently, the formal diagnosis of AD dementia, as stated by the National Institute on Aging and Alzheimer’s Association (NIA-AA), relies on neuropsychological tests further confirmed by brain imaging and cerebrospinal fluid (CSF) sampling (Jack et al., 2018).

For more than 15 years, different disease-modifying treatments (DMTs) have been investigated, yet none of them were found to be clinically effective (Buttini et al., 2005; Doody et al., 2014; Salloway et al., 2014) except for aducanumab, which was recently approved by the FDA for treatment of AD at the MCI stage (Biogen Inc., 2021; Rabinovici, 2021). However, earlier interventions remain unavailable, increasing the need for cost-effective screening biomarkers for early stage prevention clinical trials. The search for effective DMTs would largely benefit from stratification of high-risk participants in clinical trials by the implementation of minimally-invasive blood-based biomarkers that can reliable predict AD in preclinical stage individuals (Frisoni et al., 2017; Cummings, 2019). The pathogenic features of AD that have an onset in the preclinical phase may allow identification of such early blood-based biomarkers (Bateman et al., 2012; Dubois et al., 2016). By eliciting compensatory responses, these early pathogenic changes were found to initially prevent the increase in reactive oxygen and nitrogen species (ROS/RNS) through activation of antioxidant mechanisms. However, these compensatory antioxidant responses become inefficient throughout the AD continuum, resulting in progressive oxidative stress exacerbation (Katsel et al., 2013; Arce-Varas et al., 2017; Tonnies and Trushina, 2017; Merlo et al., 2019). This increase in oxidative stress has been described to induce redox post-translational modifications (PTMs) of the p53 protein, altering its native structure and disrupting its physiological functions (Uberti et al., 2002). Besides redox PTMs, *in vitro* experiments have shown that nanomolar concentrations of Aβ1-40 and Aβ1-42 can induce similar alterations in the tertiary structure of p53, previously described as the unfolded p53 conformational variant (U-p53) (Uberti et al., 2007; Lanni et al., 2010b; Lanni et al., 2013). Accordingly, higher levels of U-p53 have been consistently detected in peripheral cells derived from patients with AD as compared with controls (Lanni et al., 2008; Lanni et al., 2010a; Stanga et al., 2012). Recently, a novel antibody (2D3A8) was developed to detect this AD-specific U-p53 variant (herein described as “U-p53^AZ^”) (Abate et al., 2020), demonstrating its promising performance in predicting the AD risk at the preclinical and prodromal stages (Piccirella et al., 2021; Abate et al., 2020).

In the present study, we further assessed the potential of U-p53^AZ^ as a reliable and minimally-invasive blood-based biomarker to predict the onset of AD in cognitively normal (CN) or MCI individuals.

## Materials and methods

### Subjects

The plasma samples and supporting clinical information were provided by the Australian Imaging, Biomarkers and Lifestyle (AIBL) longitudinal cohort study, active since 2006 (Ellis et al., 2009; Fowler et al., 2021). In total, 482 subjects aged between 60 and 85 years who did not present specific comorbidities (uncontrolled diabetes, vascular disease, severe depression, or psychiatric illnesses) were included in this retrospective study collecting a total of 515 blood samples from the AIBL study by applying a consecutive sampling approach. Subjects were followed-up every 18 months. At each visit, the neuropsychological status and medical history were reviewed by a neuropsychologist and clinician. A clinical review panel consisting of a geriatrician, a neurologist, and a neuropsychologist, blinded to amyloid β-positron emission tomography (Aβ-PET) and biomarker status, determined the diagnosis of MCI and AD subjects. Data on the mini-mental state examination (MMSE), clinical dementia rating Scale (CDR) and amyloid brain burden (determined by PET with the labelled Pittsburgh compound B [PiB-PET], flutemetamol, florbetapir or NAV4694) were collected. Data on Aβ-amyloid burden was converted to the 100-points-based centiloid calibration scale (Klunk et al., 2015) and subjects were assigned to different amyloid categories according to the following scoring in centiloid units (CL): “*Negative*” <15 CL, “*Uncertain*” ≥15 CL and <25 CL, “*Moderate*” ≥25 CL and <50 CL, “*High*” ≥50 CL and <100 CL, “*Very high*” ≥100 CL. Individuals with missing imaging values were not included in the analyses of this study. Based on the NINCDS-ADRDA criteria, participants were classified as cognitive normal (CN), mild cognitive impaired (MCI) and symptomatic AD. Additionally, three subjects affected by other forms of dementia (OD) were included. The CN diagnostic group consisted of two subgroups: no memory complaints (NMC) and subjective memory complaints (SMC). The appropriate institutional ethics committee approved this study, which was carried out following all relevant ethical regulations (Human Research Ethics Committee, Research Governance Unit, St Vincent’s Healthcare, Australia (no. 028/06)).

### Blood processing and *APOE* alleles genotyping

According to the AIBL standard operating procedures, blood samples were handled at room temperature in EDTA collection tubes preventively supplemented with prostaglandin-E (final concentration: 33 ng/mL). Processing took place within three hours from blood withdrawal, followed by aliquoted sample storage in liquid nitrogen. Genotyping was carried out as has been previously described (Gupta et al., 2011).

### Antibody preparation

The 2D3A8 antibody, developed by conventional immunisation and hybridoma techniques, selectively binds U-p53^AZ^, detectable in samples from individuals with AD dementia. The antibody is selective for U-p53^AZ^ through recognition of the linear epitope carrying the sequence RRTEEENLRKKGEPHH (Memo and Uberti, 2018) which is found in human p53 between the amino acids 282 and 297. For antibody generation, healthy and disorder-free BALB/C aged between 6 and 8 weeks were injected 3 times weekly with 50 mg of the peptide (bovine serum albumin-conjugated sequence CRTEEENLRKKGEPHH, through the glutaraldehyde method), supplemented with Freund’s adjuvant. Subsequent immunisation was monitored by spectrophotometric measurements of the antibody titres after the third immunisation, and higher titres were achieved with additional inoculation. Upon selection of splenocytes with the highest antibody titre, fusion of those with mouse myeloma calls (SP2/O) followed, and its products were assessed by enzyme-linked immunosorbent assay (ELISA). Clones were then selected into 24-well plates and culture flasks.

### Immunoprecipitation (IP) and nanoflow electrospray ionisation tandem mass spectrometry (ESI-MS/MS)

Upon 2D3A8 generation, the antibody was initially used to identify peptides clinically relevant within the AD continuum through protein sequencing via MS/MS. Protein sequencing was performed at MyomicsDX Inc., Towson, MD, USA. IP took place in 2 rounds on high abundance protein-depleted plasma samples. 2D3A8 was initially used (10 μg/sample) followed by 10 µg/sample of a mixture of p53 specific antibodies (DO11:DO12:SAPU:KJC12, with respective volume ratios 1:1:2:2 and final concentration of 1 µg/µl) (Khoury and Bourdon, 2010; Joruiz and Bourdon, 2016). Upon enrichment, the peptides were eluted, fractionated through an Agilent 1290 Infinity II liquid chromatography (LC) system, and analysed in MS by a Thermo Scientific™ Q Exactive Mass Spectrometer. Tandem mass tag labelled peptides were analysed by MS/MS in a data-dependent approach on a Thermo Scientific™ EASY-nLC 1000™ HPLC system, coupled to a Thermo Scientific EASYSpray™ source supported by an analytical nanoflow column system. Acquisition of the survey full scan MS spectra (m/z 350−1800) was achieved with the Orbitrap with 35,000 resolution, following ion accumulation to a 3 × 106 target value, selected on predictive automated gain control based on the previous full scan. Sequential isolation of the 10 most intense multiply charged ions (z ≥ 2) followed, and these were then fragmented in the Axial Higher energy Collision-induced Dissociation (HCD) cell through normalised HCD collision energy at 30% (automatic gain control target: 1e5, maximal injection time: 400 ms, resolution: 35,000). The Proteome Discoverer 2.2 software processed automatically the MS raw files, and Xtract was deployed in addition to default spectrum selector node. The Mascot search engine combined with Sequest HT (interfaced with different processing nodes of Proteome Discoverer 2.2) was used to address the searches. The final dataset was reprocessed through the MyProt-QuantiR (MyOmicsDx Inc.) software package, allowing the identification of the AZ 284^®^ peptide sequence as the most clinically relevant peptide in samples from individuals affected by AD.

### Sample preparation

Fractions of samples provided by the AIBL cohort (25 µL) underwent IP using the patented 2D3A8 monoclonal antibody (Memo and Uberti, 2018) (Diadem srl, Italy) coupled to Protein L magnetic beads (ThermoFisher, USA). Upon peptide enrichment, the samples were treated with trypsin (3.5 hours at 37°C followed by 0.5 hours at 57°C).

### AlzoSure^®^ Predict test: Measurement of the AZ 284^®^ peptide in patient plasma samples

U-p53^AZ^ plasma levels were assessed through measurement of the AZ 284^®^ peptide by LC coupled to electrospray ionisation (ESI)-MS/MS at ISB srl (Italy). A surveyor HPLC Thermofisher Quaternary Pump was paired to a ThermoFisher mass spectrometer. For analyte separation, a Phenomenex Kinetex PFP column (50×4.1 mm, 2.6 m) was used and the mobile phases were A (H2O-0.2 % HCOOH) and C (CH3OH). LC was performed in a binary gradient at a chromatographic flow of 0.2 mL/min: 2% of phase C was maintained for 2 minutes and raised to 40% in 3 minutes and maintained as such for 7 additional minutes. Phase C was further raised to 70% 2 minutes later, and kept at 70% for 4 minutes before resetting to the starting conditions for 5 minutes. A calibration curve was developed prior to analysis of the clinical samples, spiking a negative control of plasma with AZ 284^®^ labelled peptide covering the range between 0.02 fmol/10 µl and 4 fmol/10µl by at least 4 different points. For analytical sessions including more than 20 samples, a spiked negative control (peptide 1 fmol/10µL) was included every 10 clinical plasma samples. Peptide sequences were then analysed with a ThermoFisher Mass Spectrometer (TSQ Vantage ThermoFisher heated-ESI ion source). The ion source parameters were: multiple reaction monitoring scan; capillary temperature: 320°C; collision energy: 5eV; sheath gas flow: 30 L/min; auxiliary gas flow: 2 L/min; and sweep gas flow: 15 L/min. Peptides were quantified through the PROSAD method and SANIST was deployed for data analysis (Cristoni et al., 2003). All tests were performed while being blinded from the clinical and cognitive data.

### Statistical analyses

The diagnostic and prognostic performance of U-p53^AZ^ to predict a certain clinical disease state were assessed and compared with the performance of amyloid status (as measured by Aβ-PET imaging, calibrated CL or inferred Aβ categories as reported above). For both diagnostic and prognostic purposes, logistic regression models were developed adding patient age and *APOE* ε4 allele status to the models.

Diagnostic analyses were performed on the biomarker data and neuropsychological assessments. While the majority of the analyses used measurements defined at the baseline assessment (i.e., the sample corresponding to the visit at which U-p53^AZ^ was measured), some were derived from subsequent follow-up visits. The predictive performance was evaluated through two specific models: 1) comparing AD individuals with non-AD individuals (including subjects in the study pooled from any other diagnosis different from AD dementia) and 2) specifically comparing CN subjects with AD dementia subjects as well as MCI subjects with AD dementia subjects. In a sub-cohort of samples with available matched amyloid status data, the diagnostic performance of U-p53^AZ^ was compared to the performance of the Aβ-PET biomarker.

Prognostic analyses were carried out using samples from SMC or MCI individuals to predict the onset of AD dementia using U-p53^AZ^. Time-dependent analyses of individuals regardless of the diagnostic status were included and compared when available with information on amyloid status. The prognostic performance of U-p53^AZ^ to predict change in clinical classification at 36 and 72 months following baseline assessment was further assessed and compared with the performance of Aβ-PET (Aβ-PET status was considered positive for a CL score ≥15).

To evaluate diagnostic and prognostic performances, receiver operating characteristic (ROC) curves were generated and their corresponding area under the curve (AUC) values were calculated to determine the accuracy, sensitivity, specificity, positive (PPV) and negative predictive values (NPV). Analyses were performed in R, using the pROC and timeROC packages for time-independent and time-dependent analyses respectively R Core Team, 2020; Robin et al., 2011; Blanche et al., 2013). ROC curve analyses to determine the optimal cut-off were performed using the cutpointr package (Thiele and Hirschfeld, 2021). Where applicable, numerical values were compared using Student’s T test or ANOVA followed by Tukey’s test for evaluation of pairwise differences. Differences in follow-up time were evaluated using a non-parametric Wilcoxon rank sum test with continuity correction. Fisher’s exact test was used to evaluate categorical count data. Comparison of ROC curves was performed through the DeLong test, while NPV and PPV were defined at a prevalence rate of AD of 30% at baseline and 45% at the final diagnosis.

## Results

### Demographics and characteristics of the study cohort

In total, 515 samples were included from 482 individuals spanning the AD continuum. Three of these individuals who were initially diagnosed with AD, but afterwards with OD, were excluded due to lacking or persistently high amyloid levels resulting in a non-congruent pathology. Out of the 479 included individuals, 422 (88%) and 472 (98%) had available information on Aβ load (quantified by CL) and *APOE* ε4 allele status (through genotyping), respectively. The demographics and clinical classification at baseline as well as the clinical classification at last follow-up are shown in **Table 1**.

**Table 1.** Baseline demographics and clinical classification of the included subjects.

|  | All patients | Non-AD individuals | AD individuals |
| --- | --- | --- | --- |
| **Total number of patients, N** | 479 | 270 | 209 |
| **Clinical status at baseline, N (%)** |  |  |  |
| NMC | 74 (15%) | 72 (27%) | 2 (1%) |
| SMC | 163 (34%) | 157 (58%) | 6 (3%) |
| MCI | 98 (21%) | 38 (14%) | 60 (29%) |
| OD | 3 (1%) | 3 (1%) | 0 (0%) |
| AD | 141 (29%) | 0 (0%) | 141 (67%) |
| **Clinical status at last follow-up, N (%)** |  |  |  |
| NMC | 65 (14%) | 65 (24%) | 0 (0%) |
| SMC | 142 (30%) | 142 (53%) | 0 (0%) |
| MCI | 48 (10%) | 48 (18%) | 0 (0%) |
| OD | 15 (3%) | 15 (6%) | 0 (0%) |
| AD | 209 (44%) | 0 (0%) | 209 (100%) |
| **Follow-up, median (months)** | 22 (0-67) | 55 (34-91) | 0 (0-18)<sup>***</sup> |
| **Age, median (IQR)** | 72 (67-78) | 71 (66 – 76) | 74 (69 – 79)<sup>**</sup> |
| **Female, N (%)** | 239 (50%) | 136 (50%) | 103 (49%) |
| **APOE4, N (%)<sup>a</sup>** |  |  |  |
| Positive (%) | 217 (46%) | 88 (33%) | 129 (64%)*** |
| Negative (%) | 255 (54%) | 182 (67%) | 73 (36%) |
| [Missing] | 7 | 0 | 7 |
| **Amyloid categories - baseline, N (%)<sup>a</sup>** |  |  |  |
| Negative | 177 (42%) | 156 (61%) | 21 (13%) |
| Uncertain | 19 (5%) | 14 (5%) | 5 (3%) |
| Moderate | 35 (8%) | 26 (10%) | 9 (6%) |
| High | 97 (23%) | 46 (18%) | 51 (31%) |
| Very high | 94 (22%) | 16 (6%) | 78 (48%) |
| [Missing] | 57 | 12 | 45 |

AD: Alzheimer's disease; CN: cognitively normal; MCI: mild cognitive impairment; NMC: no memory complaints; non-AD: subjects in the study pooled from any other diagnosis different from AD dementia; OD: other dementia; SMC: subjective memory complaints.
Aβ load categories are based on centiloid units (CL): “*Negative*” <15 CL, “*Uncertain*” ≥ 15 CL and <25 CL, “*Moderate*” ≥ 25 CL and <50 CL, “*High*” ≥ 50 CL and <100 CL, “*Very high*” ≥ 100 CL.
*P*-values in the “AD individuals” column resulted from the comparison of non-AD and AD individuals.
Numerical values were compared using a standard T-test, categorical values by Fisher's Exact test; \*\* $P \leq 0.01$ ;
\*\*\* $P \leq 0.001$ .
<sup>a</sup> Percentages were calculated on the individuals included in the analyses (i.e., missing subjects were not considered in this analysis).

During the follow-up visits, a subset of participants changed their clinical classification towards MCI, AD dementia, or OD. The total number of subjects whose clinical classification status changed during the follow-up visits is shown in **Table 2**.

**Table 2.** Overview of the number of subjects changing their clinical classification between cognitively defined groups over the follow-up of the study.

|  |  | **Final clinical classification** |  |  |  |  |
| --- | --- | --- | --- | --- | --- | --- |
| **Baseline**<br>**clinical**<br>**classification** |  | **NMC** | **SMC** | **MCI** | **OD** | **AD** |
|  | **NMC**<br>**(N=74)** | 39 | 29 | 3 | 1 | 2 |
|  | **SMC**<br>**(N=163)** | 26 | 113 | 14 | 4 | 6 |
|  | **MCI**<br>**(N=98)** | 0 | 0 | 31 | 7 | 60 |
|  | **OD**<br>**(N=3)** | 0 | 0 | 0 | 3 | 0 |
|  | **AD**<br>**(N=141)** | 0 | 0 | 0 | 0 | 141 |

AD: Alzheimer's disease; CN: cognitively normal; MCI: mild cognitive impairment; NMC: no memory complaints; OD: other dementia; SMC: subjective memory complaints.

The samples from the included subjects were further used for the diagnostic and prognostic analyses as shown in **Figure 1**.

**Figure 1.**
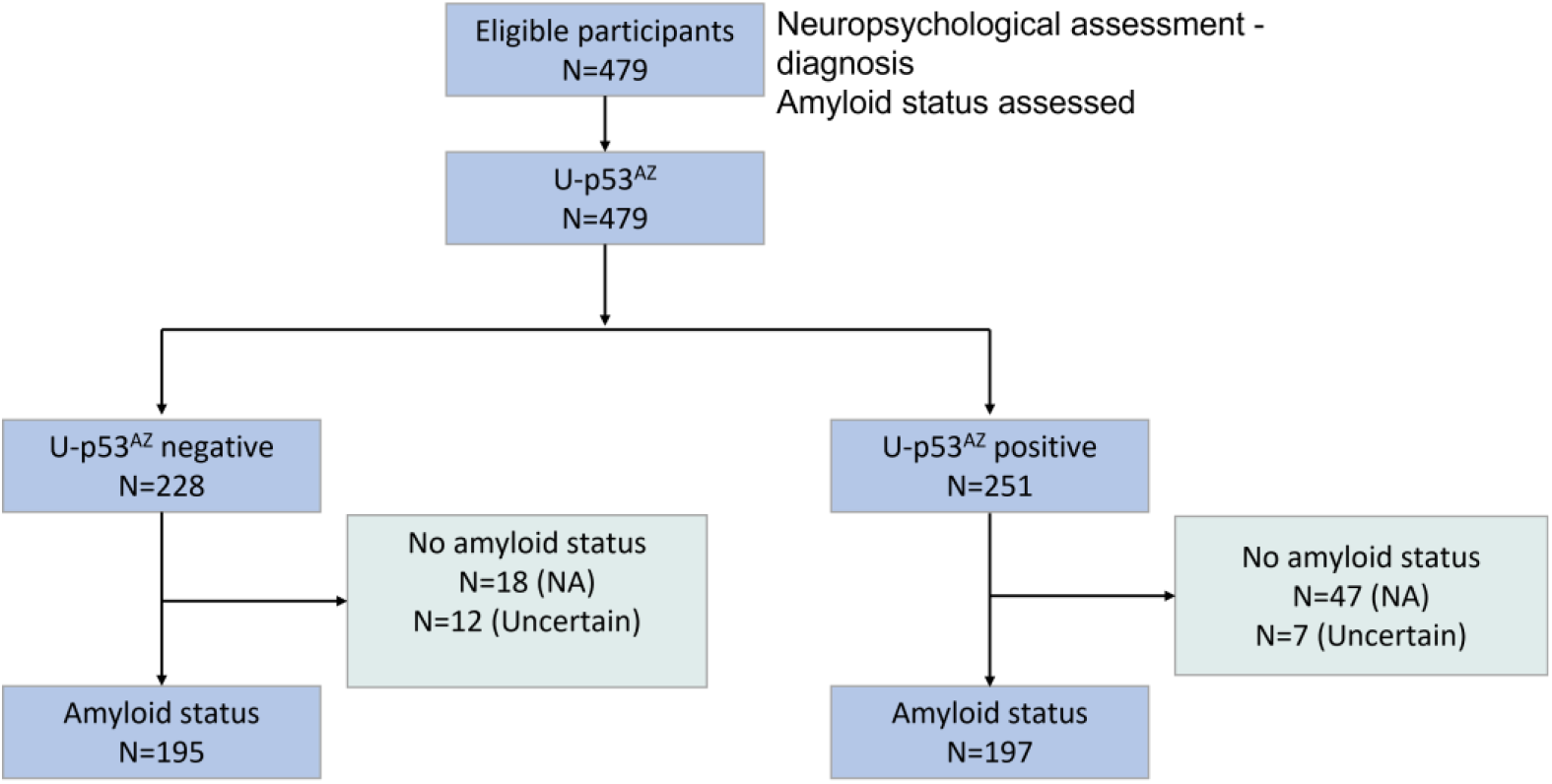
Flow of the subjects included in the study. NA: Not available.

### Performance of U-p53^AZ^ to differentiate diagnostic groups

The levels of U-p53^AZ^ were assessed relative to the clinical classifications at baseline (defined as NMC, SMC, MCI and AD dementia) (**Figure 2**). At baseline, levels of U-p53^AZ^ were similar for individuals diagnosed as NMC (N=74) and SMC (N=163) but were significantly different (ANOVA; *P*<0.001) in individuals with MCI (N=98) and AD dementia (N=141) (**Figure 2A**). Accordingly, plasma U-p53^AZ^ could accurately and reliably discriminate subjects at the preclinical stage (NMC and SMC) from those with AD (AUC=100%; 95% CI: 99-100%) with a sensitivity and specificity of both 99%. Additionally, the sensitivity and specificity of U-p53^AZ^ to differentiate subjects at the prodromal stage (MCI) from those with AD dementia (AUC=96%; 95% CI: 94-99%) were both 93%. This was confirmed at final diagnosis (i.e., last visit) with U-p53^AZ^ levels being significantly different (ANOVA; *P*<0.001) in individuals with MCI and AD (**Figure 2B**). However, levels of U-p53^AZ^ in individuals with MCI were lower at the final diagnosis as compared with the baseline diagnosis, suggesting that individuals with MCI who had high U-p53^AZ^ levels at baseline converted to AD over the follow-up of the study. Specifically, subjects (N=60) whose clinical classification changed over time from MCI to AD dementia showed significantly higher U-p53^AZ^ levels at baseline compared with subjects (N=38) who remained MCI over the follow-up of the study (*P*<0.001). Additionally, individuals who changed their clinical classification from CN to AD dementia (N=8) presented significantly higher levels of U-p53^AZ^ compared with those (N=207) who had a stable CN profile (*P*<0.01). A similar observation was noted for CN individuals (N=17) who progressed to MCI (*P*<0.001).

**Figure 2.**
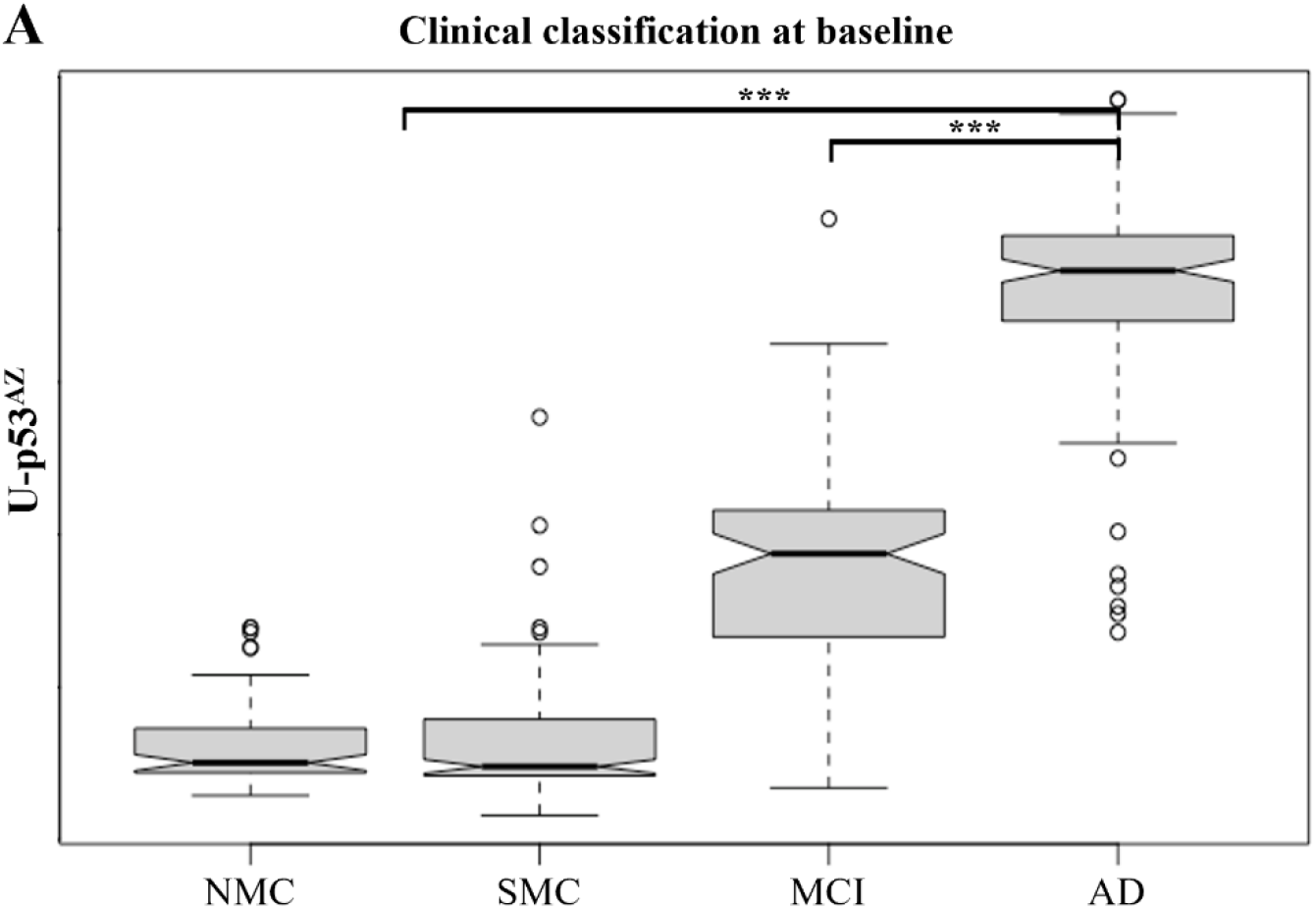
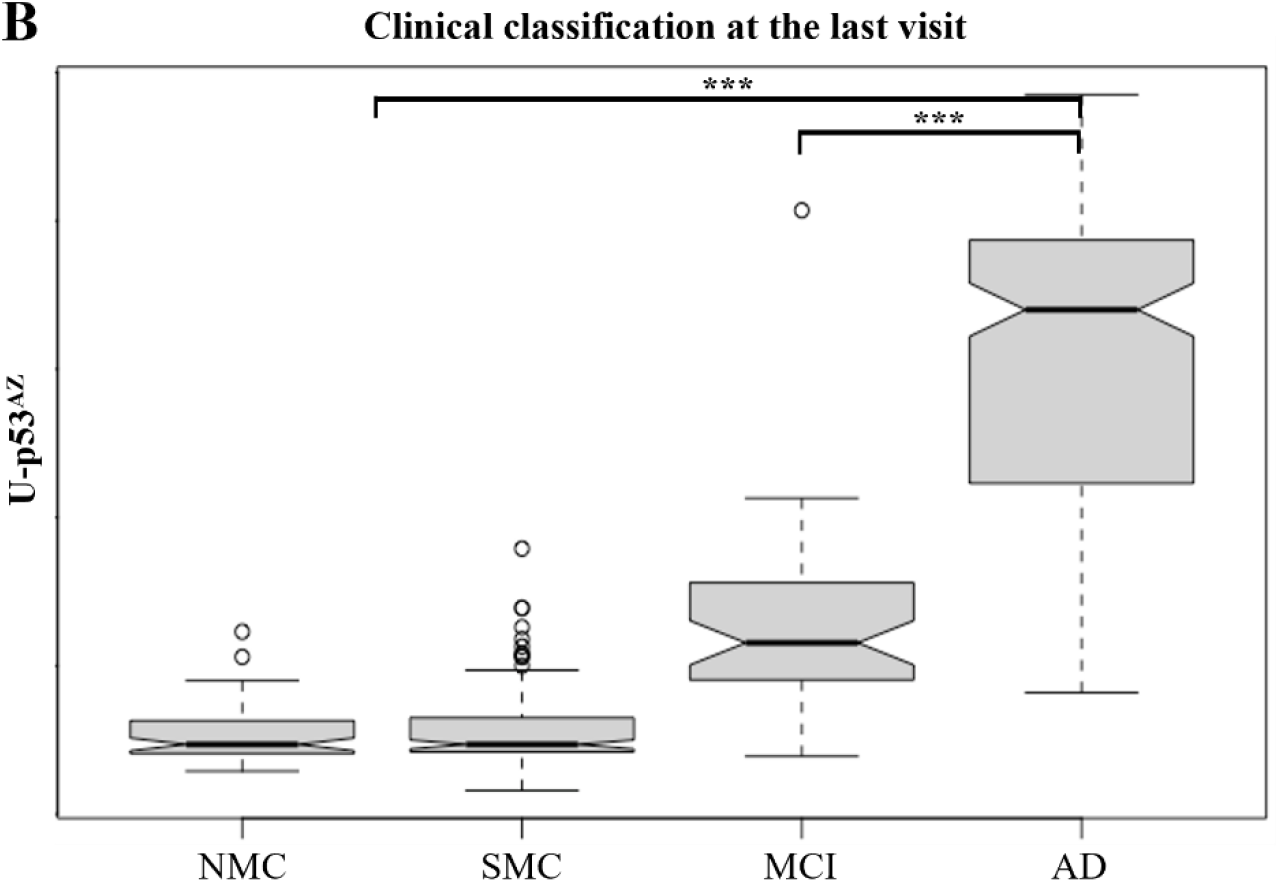
Boxplots showing the average levels of U-p53^AZ^ in different clinical classifications. A) U-p53^AZ^ levels across diagnostic groups as defined at baseline. B) U-p53^AZ^ levels across diagnostic groups as defined at the final collection (i.e., the last visit) used in this study. AD: Alzheimer’s disease; MCI: mild cognitive impairment; NMC: no memory complaints; SMC: subjective memory complaints. *** *P*<0.0001

### Diagnostic performance of U-p53^AZ^ as compared with amyloid burden

The diagnostic performance of U-p53^AZ^ to differentiate AD from non-AD individuals at baseline was significantly higher (AUC=98.6%; 95% CI:98-99%; *P*<0.0001) as compared with using amyloid status (Aβ-PET, AUC=71.9%; 95% CI:68-76%), CL numerical values (CL calibration scale; AUC=80.3%; 95% CI:76-85%) and amyloid categories based on the CL scale (AUC=78.5%; 95% CI:74-83%). Out of these three standard diagnostic approaches based on Aβ-PET, the quantitative CL (calibration scale) had the highest AUC (**Figure 3**). Hence, assessment of CL was included for further comparisons.

**Figure 3.**
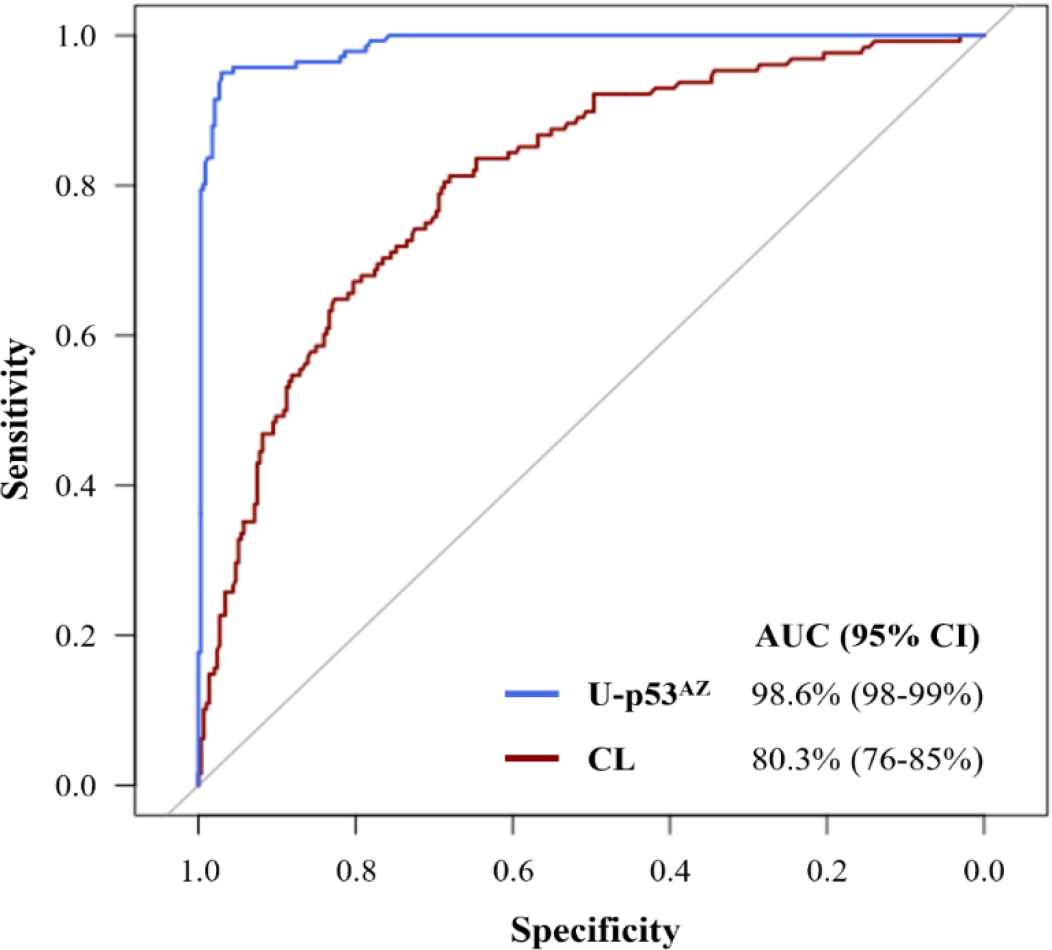
ROC curves comparing the diagnostic performance of U-p53^AZ^ versus Aβ-PET (CL numerical values) to differentiate AD from non-AD individuals at baseline diagnosis. AUC: area under curve; CI: confidence interval.

Diagnostic performance characteristics of U-p53^AZ^ and CL to predict AD dementia are reported in **Table 3**. U-p53^AZ^ was able to differentiate AD dementia from non-AD individuals with high sensitivity and specificity, outperforming Aβ-PET both at baseline and final diagnosis (*P*<0.001).

**Table 3.** Diagnostic performance of U-p53^AZ^ and Aβ-PET to discriminate AD dementia versus non-AD individuals at baseline assessment and final diagnosis.

| Metrics | Baseline |  | Final diagnosis |  |
| --- | --- | --- | --- | --- |
|  | U-p53<sup>AZ</sup> | Aβ-PET | U-p53<sup>AZ</sup> | Aβ-PET |
| AUC | 98.6% | 80.3% | 98.5% | 83.5% |
| Youden's J | 0.92 | 0.49 | 0.88 | 0.55 |
| Accuracy | 96.5% | 72.0% | 93.7% | 77.0% |
| Sensitivity | 95.0% | 81.2% | 94.3% | 79.9% |
| Specificity | 97.0% | 68.0% | 93.3% | 75.2% |
| NPV | 97.9% | 89.4% | 95.2% | 82.0% |
| PPV | 93.2% | 52.1% | 92.0% | 72.5% |

AD: Alzheimer's disease; AUC: area under the curve; NPV: negative predictive value; PPV: positive predictive value.

### The potential of U-p53^AZ^ to identify individuals at risk of AD

Both U-p53^AZ^ levels and amyloid categories (as determined by Aβ-PET) were evaluated in a subset of 448 subjects to detect individuals at high risk of AD progression based on assessment of the clinical classification. Individuals were classified in five diagnostic groups: “CN”, “MCI”, CN individuals whose clinical classification changed to MCI by the end of the follow-up (“progressors to cognitive decline [CD]”), CN or MCI individuals whose clinical classification changed to AD dementia by the end of the follow-up (“progressors to AD”) and individuals with AD dementia (“AD”). **Figure 4A** shows the classification of the different diagnostic groups by U-p53^AZ^ positivity, showing that the majority of subjects belonging to the MCI, progressors to CD/AD, and AD dementia groups were U-p53^AZ^ positive. The same set of data was classified by amyloid categories as estimated within each diagnostic group (**Figure 4B**). As expected, most individuals who were CN presented low amyloid burden, while individuals with AD dementia were assigned to mostly high or very amyloid categories. For individuals with MCI and those for whom the clinical classification changed to CD/AD dementia, classification by amyloid categories was less conclusive. For each diagnostic group, individuals with an unclear outcome from amyloid brain imaging were assigned to the category “Uncertain”. Comparison of the two methods showed a higher detection rate for U-p53^AZ^ positivity, especially for subjects whose clinical classification changed to AD dementia. As an example, 97% of subjects who changed their clinical classification to AD dementia (35/36) were classified as U-p53^AZ^ positive while only 83% (30/36) were detected by using amyloid categories.

**Figure 4.**
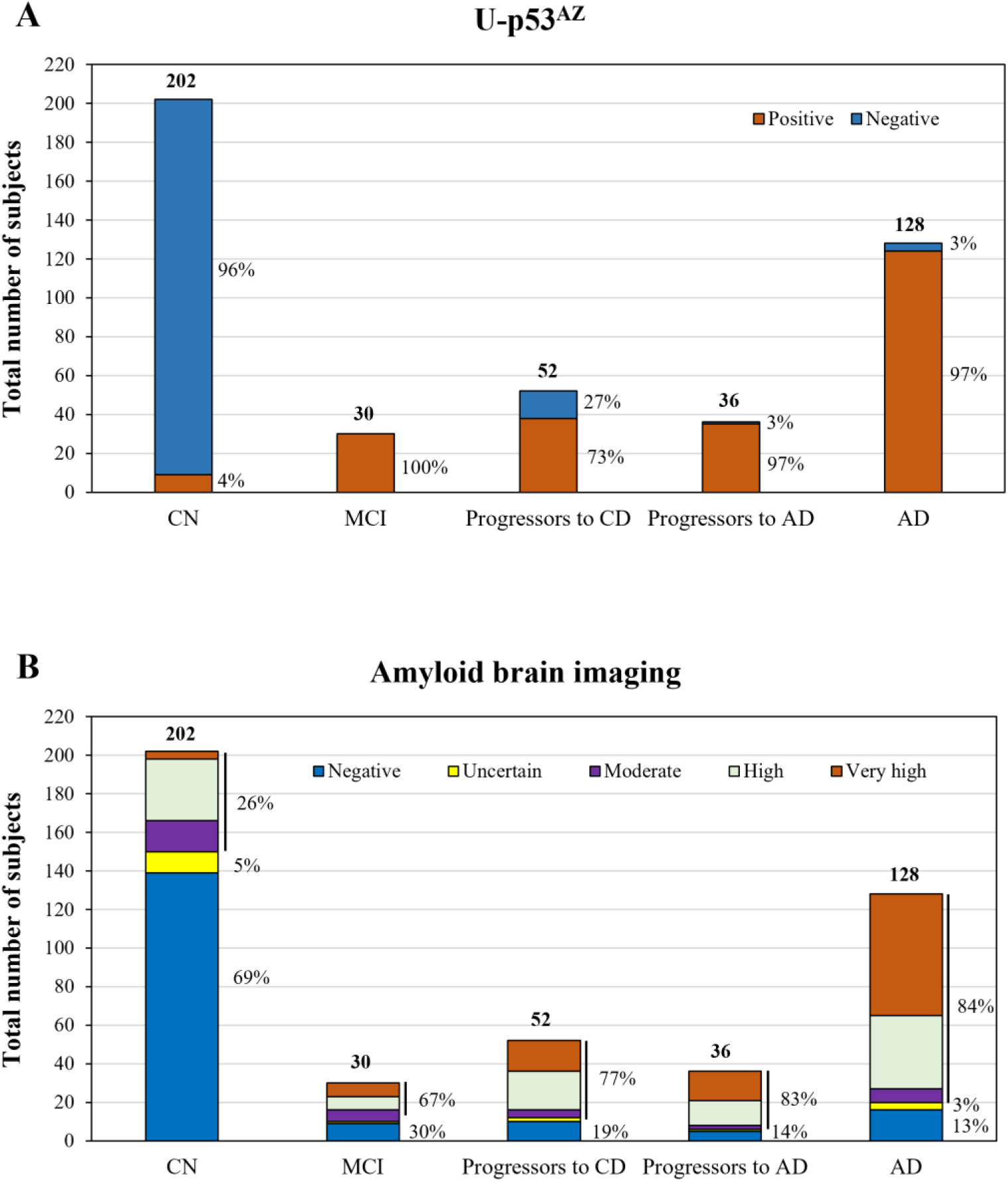
Potential of U-p53^AZ^ and Aβ categories (detected by brain imaging based on CL) to classify individuals in different diagnostic groups. A) U-p53^AZ^ positivity among clinical classifications. B) Distribution of Aβ categories based on CL among clinical classifications. AD: Alzheimer’s disease; CD: cognitive decline; CN: cognitively normal; MCI: mild cognitive impairment; NMC: no memory complaints; SMC: subjective memory complaints. In 3B, for each clinical classification the cumulative percentage of the amyloid categories “Moderate”, “High” and “Very high” is shown in place of the individual ones.

### Prognostic performance of U-p53^AZ^ to predict the onset of AD dementia

To determine the prognostic value of U-p53^AZ^, its baseline levels were assessed to predict AD dementia at different timepoints following plasma collection, independently of the initial clinical classification. In the prognostic analyses, some non-AD subjects were excluded in case insufficient data was available from follow-up visits (“censored” subjects). Based on the time-dependent ROC curves, the prognostic performance of U-p53^AZ^ was superior to amyloid brain imaging by CL at 36 and 72 following baseline assessment (**Figure 5A**). Additionally, the prognostic performance of U-p53^AZ^ was high and stable to predict the onset of AD dementia from those that were either CN or MCI at baseline, maintaining AUC values around 99% over the follow-up of the study. However, the prognostic performance of CL was lower, especially at shorter follow-up times (**Figure 5B**). The corresponding AUC values of both markers are displayed in **Table 4** confirming that the prognostic performance of U-p53^AZ^ was significantly greater than CL (*P*<0.001).

**Table 4.** Time-dependent prognostic performance of U-p53^AZ^ and CL to predict AD dementia from non-AD participants.

| Timepoint, months | 36 | 54 | 72 | 90 |
| --- | --- | --- | --- | --- |
| AD cases, N |  |  |  |  |
| U-p53<sup>AZ</sup> | 194 | 201 | 203 | 205 |
| Matched CL | 154 | 158 | 159 | 161 |
| **Non-AD cases, N** |  |  |  |  |
| U-p53<sup>AZ</sup> | 210 | 150 | 112 | 75 |
| Matched CL | 196 | 140 | 106 | 69 |
| **Censored non-AD cases, N** |  |  |  |  |
| U-p53<sup>AZ</sup> | 75 | 128 | 164 | 199 |
| Matched CL | 72 | 124 | 157 | 192 |
| **AUC (95% CI)** |  |  |  |  |
| U-p53<sup>AZ</sup> | 98% (97-99%) | 99% (99-100%) | 99% (99-100%) | 99% (98-100%) |
| Matched CL | 87% (83-91%) | 90% (86-94%) | 91% (88-95%) | 93% (89-96%) |
| *P*-value | <0.001 | <0.001 | <0.001 | <0.001 |

AD: Alzheimer's disease; AUC: area under the curve.
AUC values were compared by the DeLong test.

**Figure 5.**
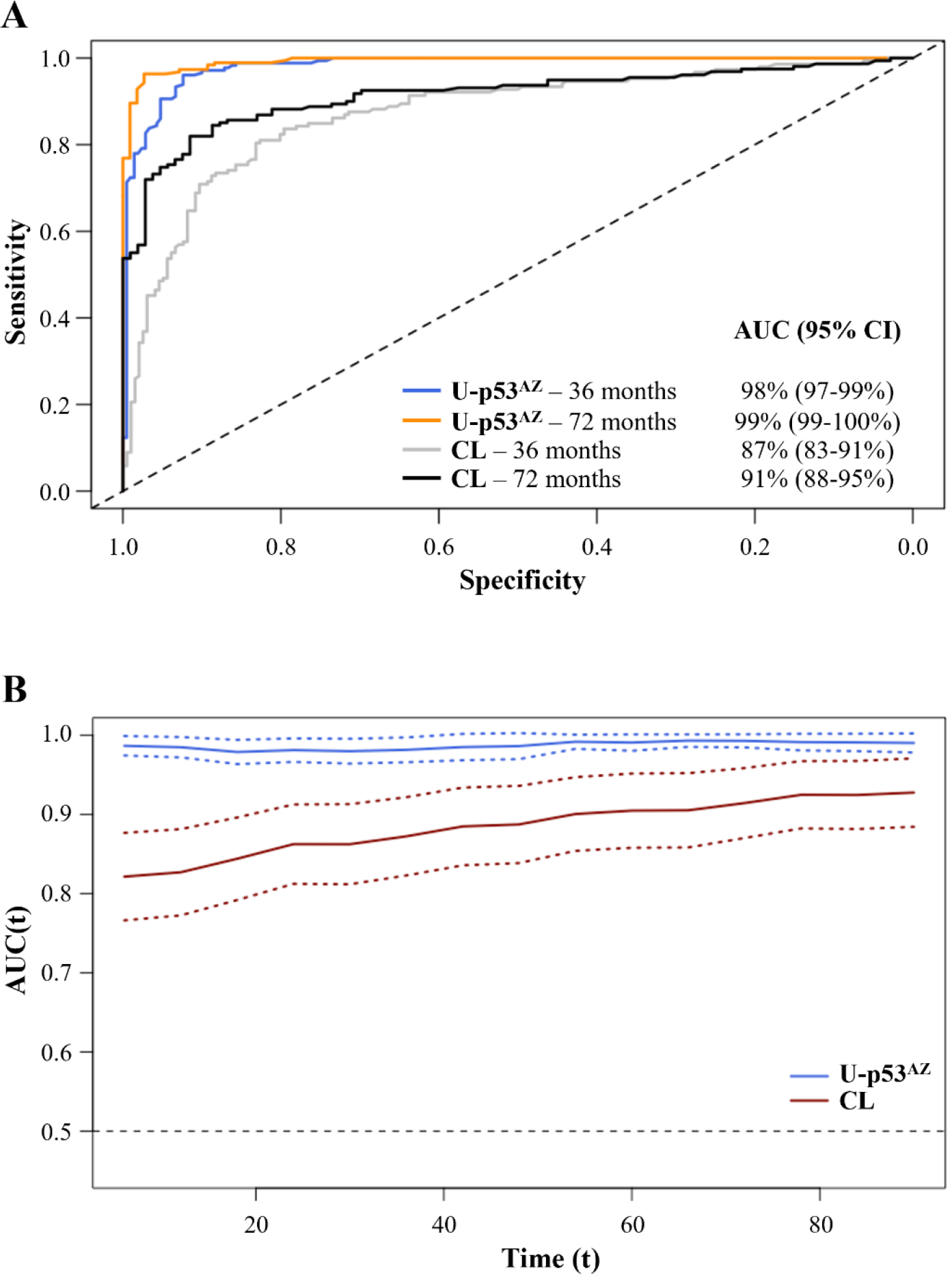
Prognostic performance of U-p53^AZ^ and Aβ categories (detected by brain imaging based on CL) in predicting the change of clinical classification to AD dementia. A) ROC curves showing the prognostic performance of U-p53^AZ^ and CL in predicting the onset of AD dementia from non-AD participants at 36 and 72 months following baseline assessment (see values in **Table 4**). B) Time-dependent AUC values (solid lines) and 95% CIs (dashed lines) of U-p53^AZ^ and CL. AUC values are calculated between month 6 and 90, in 6 months intervals. AUC: area under the curve.

Time-independent analyses similarly showed that the prognostic potential of U-p53^AZ^ in predicting the onset of AD dementia in participants with SMC or MCI to AD was high. U-p53^AZ^ predicted onset of AD dementia from participants with SMC with an AUC of 98% corresponding to a sensitivity of 100% and a specificity of 95%. The prognostic performance of U-p53^AZ^ to predict onset of AD dementia from MCI was slightly lower but still high with an AUC of 89% corresponding to a sensitivity and specificity of 77% and 93%, respectively.

### The diagnostic and prognostic performance of U-p53^AZ^ in comparison with risk factors and/or amyloid burden

Logistic regression models were used to determine the diagnostic and prognostic performance of U-p53^AZ^ in comparison with CL and other well-known risk factors of AD (i.e., age and *APOE* ε4 status) (**Table 5**). Additionally, age and *APOE* ε4 allele status were added to the U-p53^AZ^ model either alone or together with the CL model to evaluate whether inclusion of these risk factors would further increase the performance of U-p53^AZ^ in predicting the clinical classification of AD dementia and the possible change to of classification to AD dementia (**Table 5**). The diagnostic and prognostic performance of U-p53^AZ^ was superior to models based on age, *APOE* ε4 allele status and CL. As the AUC values of U-p53^AZ^ alone were nearly 100% at both follow-up times, addition of other risk factors could not further increase its diagnostic and prognostic performance, resulting in similar AUC values.

**Table 5.**
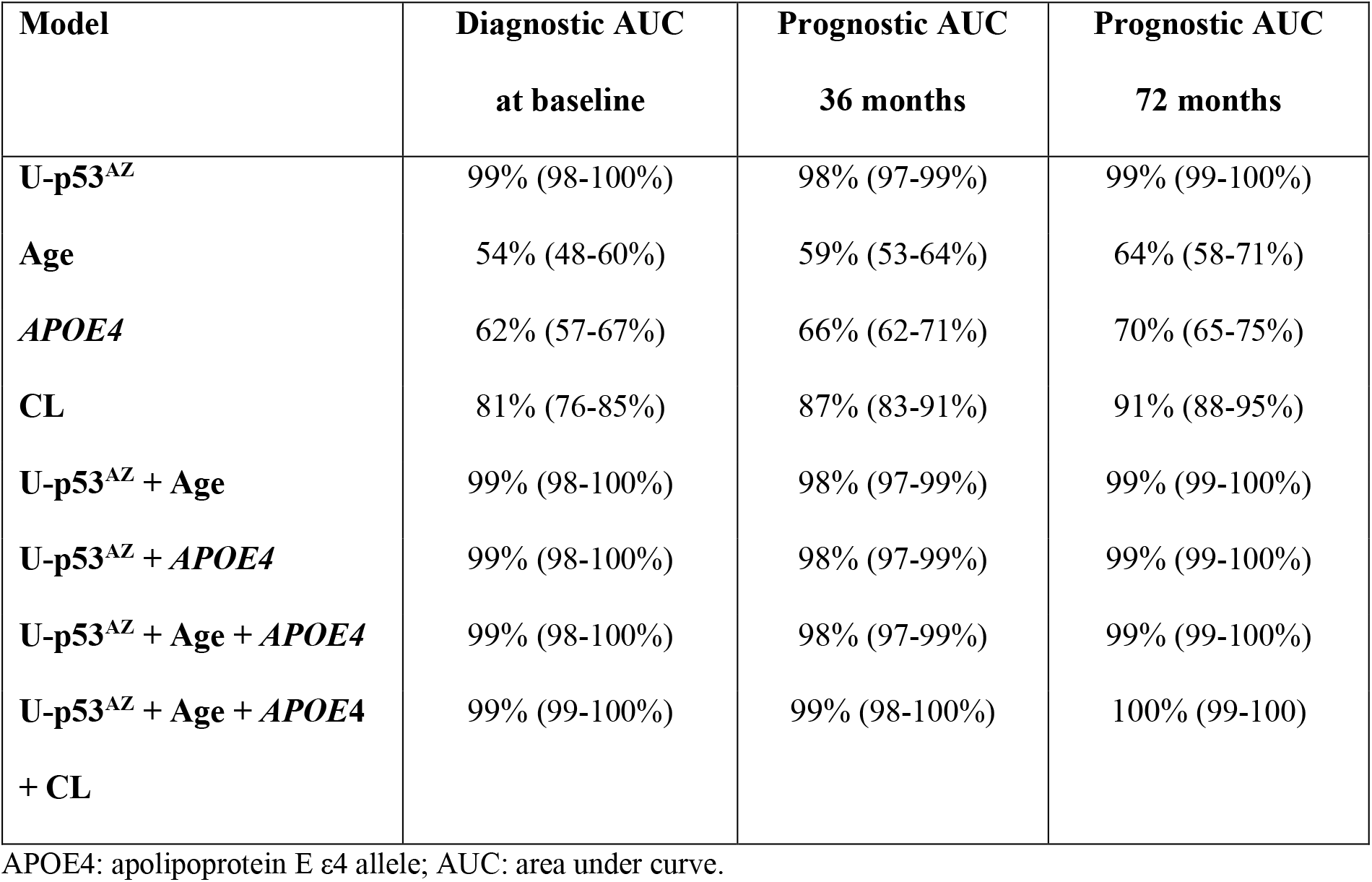
Logistic regression models showing the diagnostic and prognostic performance of different risk factors, including U-p53^AZ^ detected by U-p53^AZ^, and combinations thereof.

| **Model** | **Diagnostic AUC**<br>**at baseline** | **Prognostic AUC**<br>**36 months** | **Prognostic AUC**<br>**72 months** |
| --- | --- | --- | --- |
| **U-p53<sup>AZ</sup>** | 99% (98-100%) | 98% (97-99%) | 99% (99-100%) |
| **Age** | 54% (48-60%) | 59% (53-64%) | 64% (58-71%) |
| ***APOE4*** | 62% (57-67%) | 66% (62-71%) | 70% (65-75%) |
| **CL** | 81% (76-85%) | 87% (83-91%) | 91% (88-95%) |
| **U-p53<sup>AZ</sup> + Age** | 99% (98-100%) | 98% (97-99%) | 99% (99-100%) |
| **U-p53<sup>AZ</sup> + *APOE4*** | 99% (98-100%) | 98% (97-99%) | 99% (99-100%) |
| **U-p53<sup>AZ</sup> + Age + *APOE4*** | 99% (98-100%) | 98% (97-99%) | 99% (99-100%) |
| **U-p53<sup>AZ</sup> + Age + *APOE4***<br>**+ CL** | 99% (99-100%) | 99% (98-100%) | 100% (99-100) |

*APOE4*: apolipoprotein E ε4 allele; AUC: area under curve.

## Discussion

In the last years, there has been a growing interest in blood-based biomarkers allowing early identification of individuals at risk for developing AD (Frisoni et al., 2017). Despite these efforts, none of the studied blood-based biomarkers have yet been included in the screening protocols for the assessment of AD risk (Jack et al., 2018), leaving more than half of individuals with AD dementia without a formal diagnosis (Frisoni et al., 2017). Additionally, clinical intervention trials suffer from the lack of accessible prognostic biomarkers, leading to inadequate enrolment of individuals at risk of AD. Implementation of minimally-invasive and reliable blood-based biomarkers could respond to these unmet needs increasing the accuracy of AD diagnosis while improving clinical trial enrolment (Frisoni et al., 2017; Cummings, 2019). This is particularly relevant in the coming years due to aging of the population and the subsequent increase in the numbers of patients receiving the clinical diagnosis of AD (Horgan et al., 2020).

One of the recently discovered blood-based biomarker candidates is a conformational variant of p53. Although p53 is mostly known for its role as tumour suppressor, it is now clear that p53 is regulating cell fate decisions and preventing neurodegeneration through multiple mechanisms, including DNA damage repair, protection from oxidative stress, support of axonal regeneration, neuronal outgrowth and synaptic function as well as repression of β-site amyloid precursor protein-cleaving enzyme 1 (BACE1) expression (Di Giovanni et al., 2006; Tedeschi et al., 2009; Merlo et al., 2014; Silva et al., 2014; Singh and Pati, 2015; Labuschagne et al., 2018). Nevertheless, its neuroprotective function becomes gradually lost throughout the AD continuum, as exposure to increasing amounts of amyloid beta and ROS/RNS leads to unfolding of the p53 protein resulting in elevated levels of the p53 conformational variant, identified as U-p53 (Uberti et al., 2002; Cenini et al., 2008; Lanni et al., 2010b; Lanni et al., 2013). U-p53 has previously been detected in peripheral cells derived from MCI and AD individuals and was shown to be predictive of the onset of AD dementia (Lanni et al., 2008; Lanni et al., 2010a; Stanga et al., 2012). Although promising, these preliminary studies were based on limited patient numbers and conventional techniques, such as flow cytometry and ELISA, using commercially available antibodies (Lanni et al., 2010a; Stanga et al., 2012). Assessment of U-p53 has now been improved by the development of a novel antibody (2D3A8) specifically targeting the AD clinically relevant U-p53 conformational variant (U-p53^AZ^) (Piccirella et al., 2021; Abate et al., 2020). To further improve its analytical performance, U-p53^AZ^ was quantified by the AlzoSure^®^ Predict method, a LC-ESI/MS/MS sequential technique using plasma samples from individuals at different stages of the AD continuum, available through the longitudinal AIBL cohort. Due to its high specificity in comparison with conventional techniques and its increasing accessibility at lower costs, MS has been widely used in several blood-based biomarker studies (Jannetto and Fitzgerald, 2016; Oeckl and Otto, 2019; Stevens and Pukala, 2020). By using this technique, the present study aimed to assess whether quantification of U-p53^AZ^ was able to assign individuals to the correct diagnostic group and determine its performance for predicting the onset of AD at the preclinical and prodromal stages.

This study showed that baseline U-p53^AZ^ levels were highest in individuals with AD allowing accurate differentiation of individuals in the preclinical and prodromal stages from those with AD dementia (AUC values ≥97%). Similarly, U-p53^AZ^ could more accurately detect AD dementia onset compared to Aβ-PET showing its potential to identify individuals at high risk to develop AD dementia. Furthermore, U-p53^AZ^ was able to differentiate AD individuals from non-AD ones up to 6 years prior to AD dementia onset with an AUC of 99%, again performing significantly better than Aβ-PET. Additionally, the predictive performance of U-p53^AZ^ markedly surpassed those of other risk factors (dichotomous *APOE* ε4 allele status, Aβ-PET, age) used either alone or in different combinations. Generally, the herein presented results confirm the ones from a previously published study using a similar technique. However, by including a larger study cohort the predictive performance of U-p53^AZ^ was found to be further improved (Piccirella et al., 2021). When comparing to a model based on magnetic resonance imaging (MRI), PET, CSF and covariates (Shaffer et al., 2013), the predictive potential of U-p53^AZ^ was higher and without the limitations associated with costs and invasiveness.

While the development of blood-based biomarkers was initially intended for diagnostic purposes, their potential in accurate prediction of the onset of AD in non-demented CN adults and MCI patients is gaining more interest (Shen et al., 2020). As an example, plasma phospho-tau181 levels were recently shown (Janelidze et al., 2020) to predict the change of clinical classification of individuals from SMC to clinical AD dementia with AUC values comparable to U-p53^AZ^ (Stockmann et al., 2020). This poses the question whether screening based on an unique biomarker instead of a panel could be used for the precise identification of individuals who are prone to develop AD dementia. Further comparisons of the predictive performance of U-p53^AZ^ with Aβ-PET and other clinical markers, such as tau, are needed to validate the potential role of U-p53^AZ^ as unique prognostic blood-based biomarker.

Although our findings are promising and suggestive of the predictive potential of U-p53^AZ^, the results herein described are based on a single longitudinal study. Further studies should include larger sample numbers provided by different centres to validate the performance of U-p53^AZ^ in predicting onset of AD dementia and to determine a cut-off that can be integrated in clinical practice for stratification of CN individuals who are at high risk to develop AD dementia.

Currently, we are conducting a follow-up study using additional longitudinal data from different cohorts and centres including individuals from Europe and USA to validate the herein presented findings and to compare and correlate the potential of U-p53^AZ^ as blood-based biomarker with traditionally studied markers of AD pathology (e.g., tau deposits). Additionally, assuming that preventing the development of AD dementia could result in the reduced increase of U-p53^AZ^ levels, future studies should examine if U-p53^AZ^ could support patient stratification in clinical trials and assessment of DMTs effectiveness.

In conclusion, the present study demonstrated the high performance of U-p53^AZ^ to predict the onset of AD in asymptomatic individuals or individuals at the prodromal stage as early as six years prior to a clinical AD dementia diagnosis. We believe that quantification of U-p53^AZ^ through AlzoSure^®^ Predict has the potential to be implemented in screening strategies, allowing improved stratification of individuals at high risk to develop AD dementia in clinical intervention trials to subsequently support the development of effective DMTs. Ultimately, early detection of AD could ensure timely prescription of DMTs to maximise their efficacy.

## Supporting information

STARD checklist

## Data Availability

Data available on request from the authors.

## Abbreviations

Aβ: Amyloid beta
AIBL: Australian Imaging, Biomarkers, and Lifestyle
AUC: Area under the receiver operating characteristic curve
AD: Alzheimer’s disease
APOE ε4: Apolipoprotein E ε4 allele
BACE1: β-site amyloid precursor protein-cleaving enzyme 1
CI: Confidence interval
CN: Cognitively normal
CSF: Cerebrospinal fluid
ELISA: Enzyme linked immunosorbent assay
ESI: Electrospray ionisation
HCD: Higher energy collision-induced dissociation
HPLC: High performance liquid chromatography
IP: Immunoprecipitation
MCI: Mild cognitive impairment
MRI: Magnetic resonance imaging
MS/MS: Tandem mass spectrometry
OD: Other types of dementias
PiB-PET: Positron emission tomography utilising Pittsburgh compound B
ROC: Receiver operating characteristic
PTMs: Post-translational modifications
RNS/ROS: Reactive nitrogen/reactive oxygen species
SACI: Surface-activated chemical ionisation
U-p53: unfolded p53

## Authors’ contribution

SP with PK developed the study design, SP overviewed the development of the protocol. LVN and CX contributed to the biostatistics analyses. All authors contributed equally to the manuscript. All authors read and approved the final manuscript.

## Ethics approval and consent to participate

An appropriate institutional ethics committee approved the AIBL study, which followed relevant ethical regulations. Further information: Human Research Ethics Committee, Research Governance Unit, St Vincent’s Healthcare, Australia (no. 028/06). Individuals registered in AIBL and could opt out of PiB-PET. Their plasma samples are stored in the biobank and a subset of them was requested for this exploratory study.

## Consent for publication

All authors agreed to the manuscript prior to submission.

## Availability of data and materials

Data available on request from the authors

## Acknowledgments

We would like to thank Simone Cristoni from ISB for MS consultancy and testing and Ismar Healthcare NV for the medical writing assistance in support of Diadem srl, Brescia, Italy.

